# Prediction of the Epidemic Peak of Covid19 in Egypt, 2020

**DOI:** 10.1101/2020.04.30.20086751

**Authors:** Eman D. El Desouky

**Author notes:** **Address:** Kasr Al Eini ‘Street ‘Fom El Khalig ‘Cairo Governorate, **Postal code:** 11796.

## Abstract

**Objectives:** Since December 2019 a pandemic of new novel coronavirus has started from Wuhan, China, in Egypt, the first case reported on February 14, 2020. In this study we aimed to predict the time of possible peak and simulate the changes could be happen by the social behavior of Egyptians during Ramadan (the holy month).

**Methods:** SIR and SEIR compartmental models were used to predict the peak time. We simulated different expected scenarios based to examine their effects on the peak timing.

**Results:** We found that the peak most likely to be in middle of June 2020. Simulating different transmission rate probability and R0 the earliest peak could to be in the May 20 and latest one could be in 18 July. The peak shifted much earlier to 11^th^ April 2020 without lockdown and other mitigation strategies.

**Conclusion:** Social behaviors of citizens during the holy month will dramatically affect the peak timing. Mitigations strategies and other lockdown measure helped to delay the expected peak.

## Introduction

The first case of respiratory disease caused by a novel coronavirus was identified in Wuhan City, Hubei Province, China in In December 2019 and on 13 January 2020 the first case outside of China was reported in Thailand.^1^ On 11^th^ March 2020 the World Health Organization named it coronavirus disease 2019 (COVID-19) and considered as pandemic.^2^ The first case appeared in Egypt on 14^th^ February 2020 for a foreigner and fortunately was asymptomatic until 1^st^ of March 2020 he was the only reported case after that few cases started to appear till 14^th^ March 2020 to reach 110 cumulatively, and then the number of laboratory confirmed cases increased weekly (Table 1) to reach 3490 till April 22, 2020.^3^

**Table 1.**
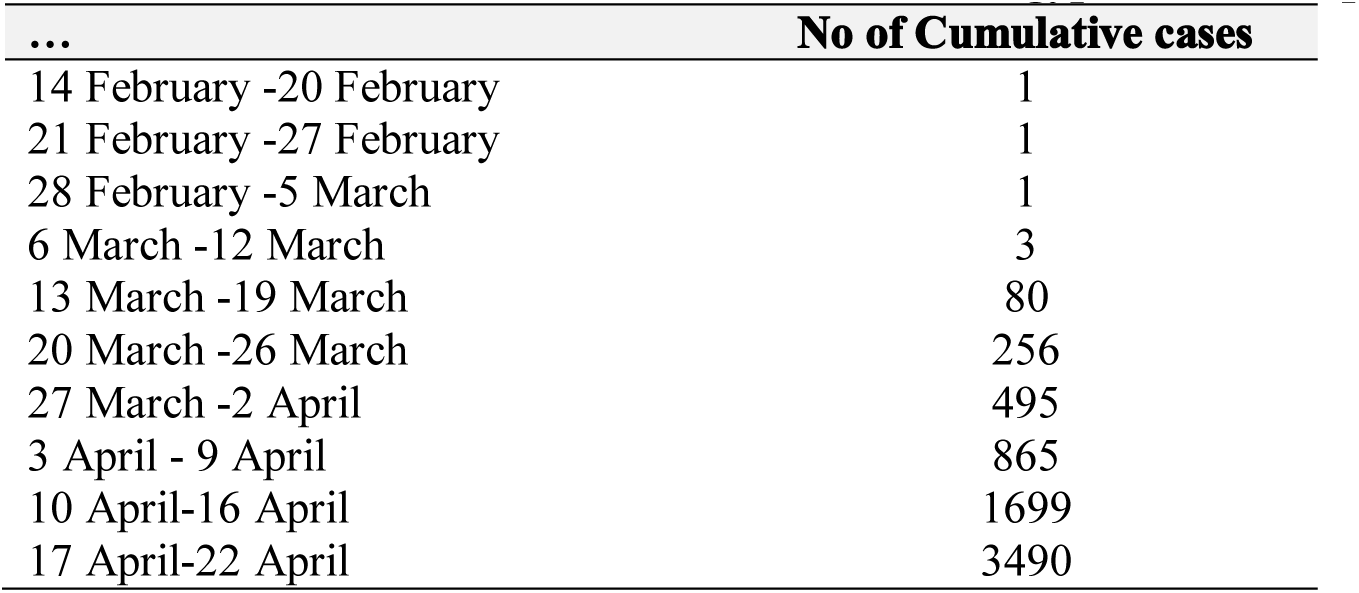
Number of cumulative COVID-19 cases in Egypt until 23^th^ April 2020.

As seen in table 1 the numbers started nearly to double weekly from the fifth week and after closure of schools with partial lockdown the public concerned about the timing of the cases surge. The aim of this study is to predict the time of possible peak and simulate the changes that might happen by the social behavior of Egyptians during Ramadan (the holy month) that will begins on Friday April 24 and ends on Saturday May 23. In Ramadan families usually gather especially in first days, children have more activates with each other’s and everyone leaves work at the same time and need to be in home at the same time; currently with Covid19 pandemic and partial lock down it’s challenging to cope with social distancing and Ramadan habits.

## Methods

We tested two models SIR and SEIR models ^4^ to predict peak timing using data till 17th April 2020 and simulated different scenarios based on hypothetical expected changes in social behavior of Egyptians during Ramadan.

### 1- SIR model

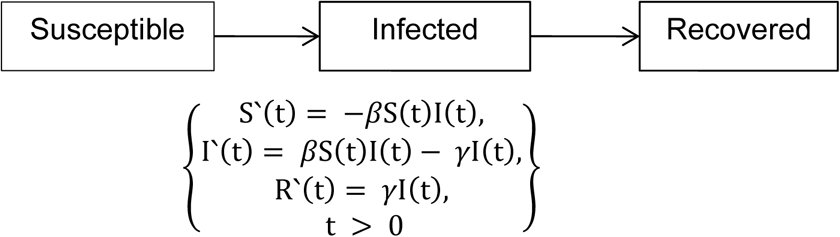

### 2- SEIR model

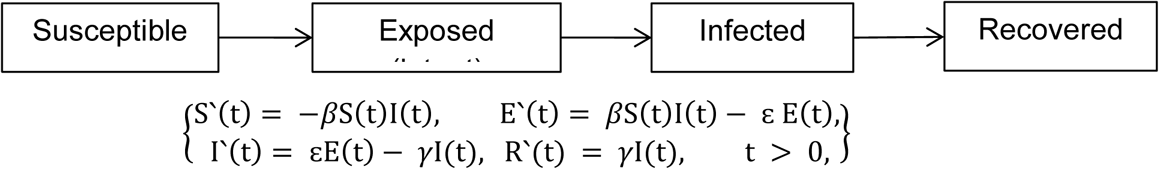

Where S(t), E(t), I(t) and R(t) represent the susceptible, exposed, infective and removed populations at time t, respectively. β, ε and γ represent the infection rate, the onset rate and the removal rate, respectively. 1/ε and 1/γ imply the average incubation period and the average infectious period, respectively. The force of infection (rate of infection per unit time is βI and β denoted the rate of transmission from S to I or effective contact between S and I.^5^

These models based on Euler’s Method: A numerical method for solving differential equations and referred to as a compartment model since it is useful to refer to people moving from one compartment to another.

Parameters used in the models based on previous studies assuming average Incubation period = 5 days and so onset rate ε= 0.2 ^6^, assuming infectious period = 10 day and so removal rate γ=0.1.^7^ Egypt population (1×10^8^) ^8^, no one infected at the time (t_0_) time step is one day and population remain constant so S±E+I+R=1.

Regarding transmission rate (β) and Basic reproduction number (R_0_) researcher depended on the published data available on GitHub, midas-network^9^, R_0_ for Egypt as published 2.3 and so β =0.23 by applying the formula;^10^

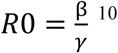

For the purpose of simulation different scenario of effective contacts during Ramadan, different R_0_ and β were used within the range of previous published data available on midas-network.^9^

Starting from model1 (M1) that represent less effective contact rate than the current situation (β =0.22, R_0_=2.2) till model10 (M10) that represent higher effective contact rate with β =0.32 and R_0_=3.2

The peak was identified by the maximum proportion of population infected in a year

## Results

Researcher used SIR model with different simulation scenarios to predict peak timing, illustrated in figure 1. The earliest of peak could be on May 20, 2020 and latest peak could be July 18, 2020, then applied Applying regression method to estimate the best fitted models with the cumulative data released by MOHP till 17th April, 2020; peak on 15 June 2020 is more likely.

**Figure (1):**
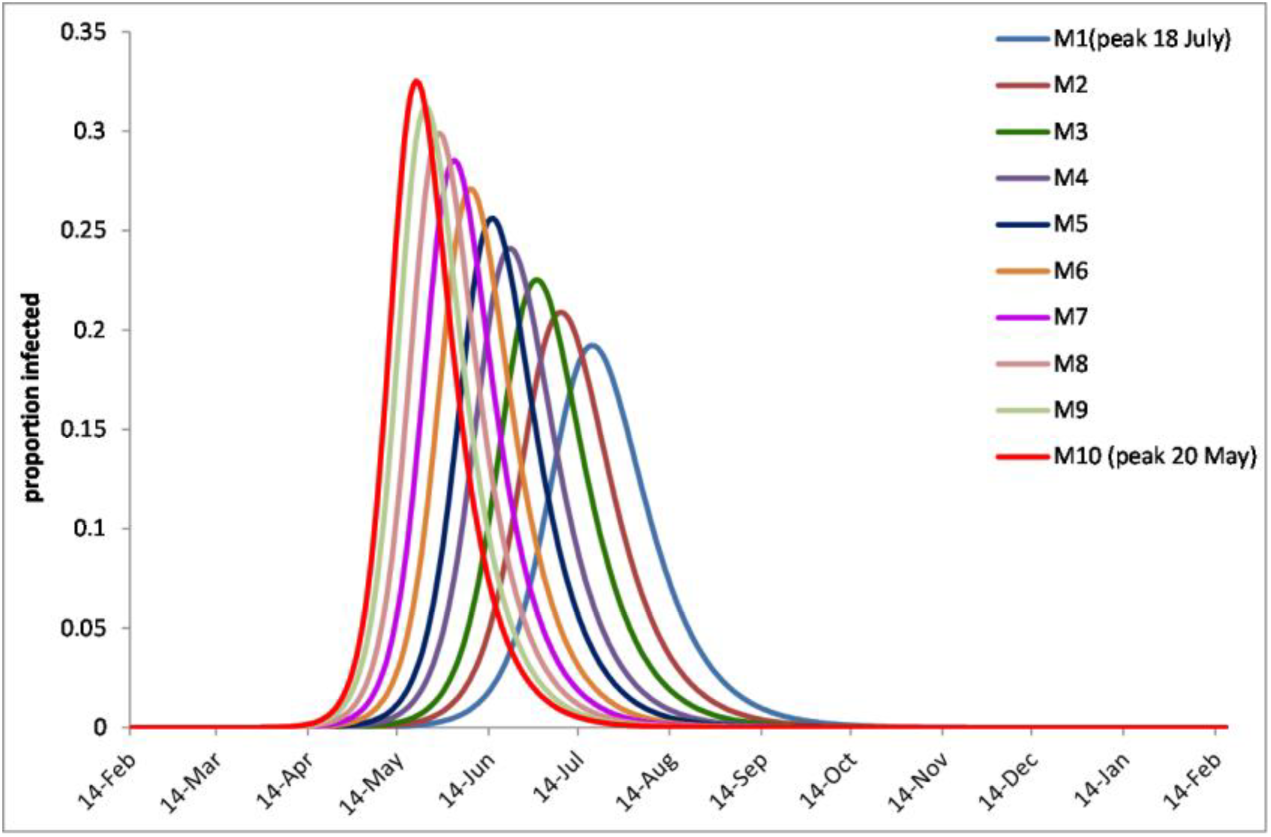
Different models estimating different peaks with SIR model

Researcher used SEIR Peak prediction models almost similar results obtained; earliest peak could be May 17, 2020 and latest peak could be by July 15, 2020.

The 3^rd^ scenario was estimating the peak time without lockdown and other strategies based on mobility changes report released by google^11^ that stated average 50% reduction in Egypt since the time of lockdown and mitigation strategies. The peak was estimated to occur on 11^th^ April 2020 but with mitigation strategies and lockdown it was shifted about 69 days to reach June 15, 2020 (figure 2).

**Figure (2):**
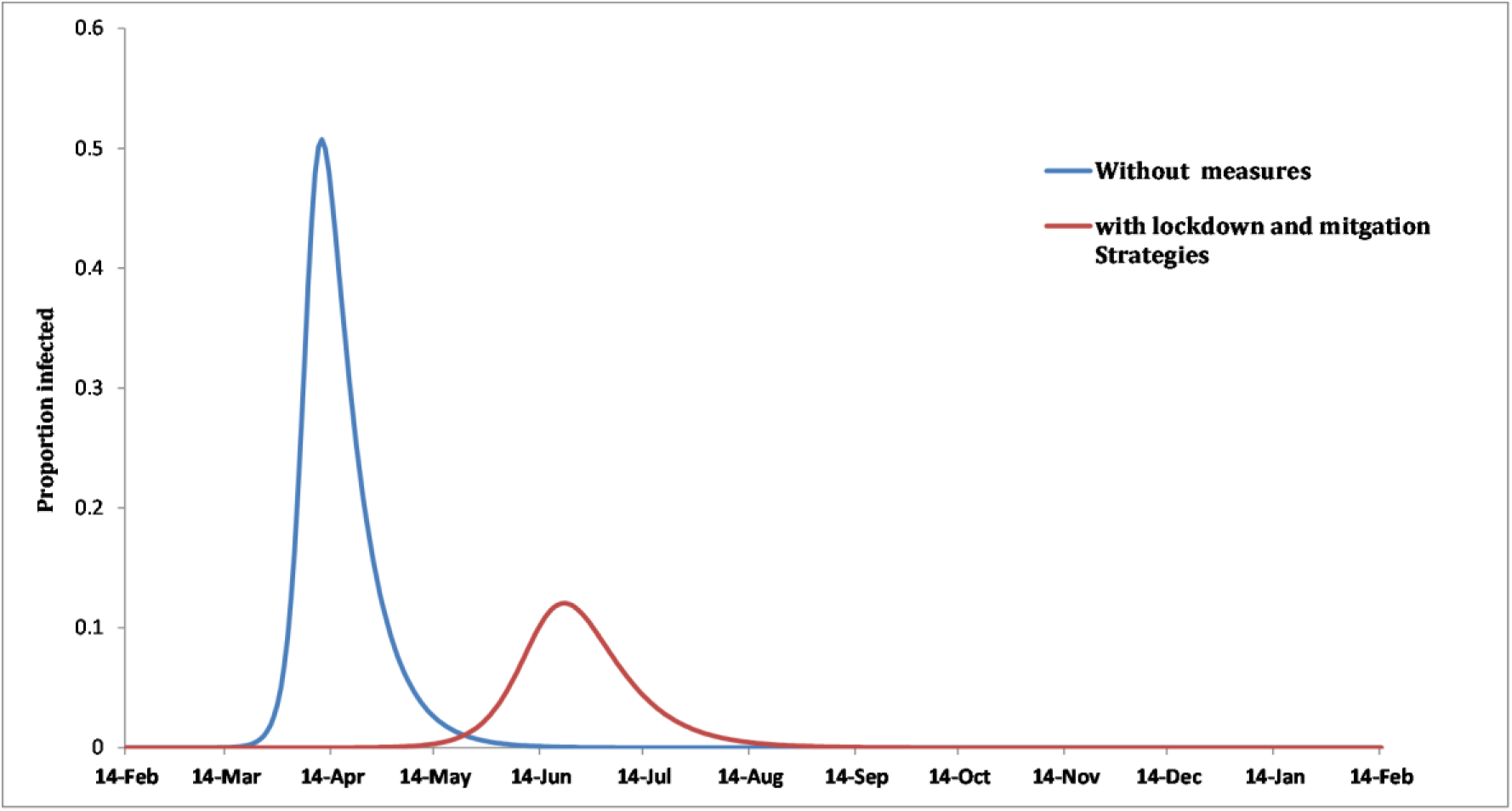
Peak estimation with lockdown and other measures in comparison to the situation without them.

## Discussion

In this study, by applying the SIR and SEIR compartmental model along with the daily reported cases of Covid19 till 17^th^ April in Egypt; we estimated the peak could be in the middle of June. We hypothesized that the social and contact rate per individual changes in Ramadan (the Holy month) that will start on April 24, 2020 will affect the timing of peak as a result of Increase in β (transmission rate). We simulated different situations with different rates of effective contact, transmission rate and R_0_ consequently; the peak timing changed to reach 20 May, 2020 as earliest one and latest peak could be July 18, 2020. These findings was in concordance with the study done by **Pearsn et al** ^12^ that reported 10,000 cases could be in 20 May 2020. On the contrast a recent report released by **SUTD Data-Driven Innovation Lab**^13^ that reported the pandemic will end in Egypt on May 20, 2020. Researcher simulated another scenario based on current data to predict the timing of peak without lockdown the peak shifted earlier to be on April 11, 2020; the mitigation strategies helped to delay peak. The current study simulated different scenarios based on different values on transmission rate (calculated and estimated) in concordance with **Bootsma and Ferguson**^14^ who stated the most important factor affecting disease pandemic is the transmission rate which depend on crowding as it affects amount of contact per individual. The current study used SIR and SEIR models which are too idealistic for modeling COVID-19 through numerical simulation, but it gives an idea about when the COVID-19 cases can surge to.^15^

The study stated long range peak prediction to emphasis the role of social distancing to flatten the curve in concordance with **Anderson et al**^16^.

The result suggests that the epidemic of COVID-19 in Egypt would not end so quickly. This might be reliable with the WHO’s statement on 6 March 2020 that it is a false hope that COVID-19 will disappear in the summer like the flu. ^17^

## Limitations

This is too idealistic for modeling COVID-19 as the reproduction number (R0) changes overtime but the multiple simulation may overcome this hypothetically for certain time. The deterministic models are not accurate and used to give insight about what will happen to population on average. Study didn’t estimate the hospitalized number at the surge however the timing of the peak didn’t affected by the identified cases form the actual infective population.

## Conclusions

During COVID-19 pandemic it’s hard to predict the real peak timing however the surge timing will depend mainly on the behavior of citizens towards social distancing and hygiene measures. The mitigation strategies and lockdown in Egypt has a positive effect on the delay of the epidemic peak, giving more time to the health sector to encompass the situation. There is an urgent need for Local and worldwide policy to deal with Covid19 that will extend to summer time. Egyptian government should monitor the reported cases daily along with the performance of citizens in the coming month to determine the proper strategies to flatten the curve as much as possible. The optimistic view to the situation should be treated cautiously as COVID-19 is still unclear.

## Data Availability

data will be available on request

